# Benefits of Surveillance Testing and Quarantine in a SARS-CoV-2 Vaccinated Population of Students on a University Campus

**DOI:** 10.1101/2021.06.15.21258928

**Authors:** Francis C. Motta, Kevin A. McGoff, Anastasia Deckard, Cameron R. Wolfe, M. Anthony Moody, Kyle Cavanaugh, Thomas N. Denny, John Harer, Steven B. Haase

## Abstract

Surveillance testing and quarantine have been effective measures for limiting SARS-CoV-2 transmission on university campuses. However, the importance of these measures needs to be re-evaluated in the context of a complex and rapidly changing environment that includes vaccines, variants, and waning immunity. Also, recent guidelines from the CDC suggest that vaccinated students do not need to participate in surveillance testing. We used an agent-based SEIR model to evaluate the utility of surveillance testing and quarantine in a fully vaccinated student population where vaccine effectiveness may be impacted by the type of vaccination, the presence of variants, and the loss of vaccine-induced or natural immunity over time. We found that weekly surveillance testing at 90% vaccine effectiveness only marginally reduces viral transmission as compared to no testing. However, at 50%-75% effectiveness, surveillance testing can provide over 10-fold reduction in the number of infections on campus over the course of the semester. We also show that a 10-day quarantine protocol for exposures has limited effect on infections until vaccine effectiveness drops to 50%, and that increased surveillance testing for exposures is at least as effective as quarantine at limiting infections. Together these findings provide a foundation for universities to design appropriate mitigation protocols for the 2021-2022 academic year.

## Introduction

Surveillance testing and quarantine were two approaches used successfully by universities to limit the spread of SARS-CoV-2 in the student population during the 2020-2021 academic year[2]. In the coming year, many universities are looking to vaccinations as the foundation of their COVID-19 mitigation programs, with some requiring a SARS-CoV-2 vaccination to live and attend classes on campus. If 100% of the student population is vaccinated, will surveillance testing and quarantine still be important for preventing viral transmission in the student body?

If all students were vaccinated and the vaccinations were 100% effective, then surveillance testing and quarantine would be unnecessary. However, the BNT162b2 and mRNA-1273 vaccines exhibit ∼90% effectiveness against infection [3-5], and the Ad26.COV2.S vaccine is ∼66%[6]. Moreover, some genetic variants of SARS-CoV-2 (e.g., B.1.351 (beta) and P1 (gamma)) appear to have some resistance to vaccine-induced neutralizing antibodies [7-10]. Thus, it is possible that the effectiveness of vaccines may be further eroded by the presence of known and future viral variants [11]. Finally, the durability of vaccine-induced immunity is still unknown, and it is likely that immunity will wane over time [12, 13]. All of these factors are likely to contribute to an uncertain and continually changing environment of immunity in a vaccinated population of students. In such an environment, it is conceivable that surveillance testing and quarantine might continue to have an important role to play in COVID-19 mitigation strategies on college campuses, in contrast to recent CDC guidelines [1].

Mathematical models have been useful tools for exploring SARS-CoV-2 infection dynamics and the effects of a variety of mitigations on university campuses [14-17]. Here we used an agent-based, modified SEIR model to investigate the effect of surveillance testing and quarantine in an environment where 100% of the student population is vaccinated, but where vaccine effectiveness may be reduced by variants or by waning immunity. We also sought to understand whether quarantine was still a useful method for preventing transmission and whether an increased surveillance testing protocol was as effective as quarantine at limiting viral transmission.

Unknown model parameters were estimated from infection data collected by Duke University’s surveillance testing program during the 2020-2021 academic year [2]. Although the precise model outputs will be partially tuned to Duke-specific parameters, we expect that general trends will be applicable to a variety of university campus environments.

## Methods

### SEIR Model

We used an agent-based model with SEIR disease dynamics to simulate SARS-CoV-2 spread in a population of 5000 homogeneously mixing agents, which constitutes the on-campus population and is the focus of our model. We also considered an off-campus population, which we modelled as a reservoir with a static prevalence of infection that is not influenced by the disease dynamics of the modelled population. The disease dynamics progress as follows. Each day each susceptible agent may randomly become exposed through a fixed number of interactions with both the on-campus and the off-campus populations. Each agent’s probability of exposure depends on the prevalence of infection in the (non-isolated and non-quarantined) populations with which it is interacting and on a fixed probability that an interaction will lead to an exposure.

Because the probability that an interaction will result in an exposure is highly context-specific and difficult to estimate [18-20], we arbitrarily fixed the transmission probability due to an interaction, and thereby implicitly defined the type of interactions being modelled via this probability. We then estimated reasonable numbers of such interactions by simultaneously searching for the number of daily between-agent interactions and the number of daily interactions between each agent and the outside community that minimized the error between the model output of daily positive cases and the daily positive cases at Duke University during the Spring 2021 semester. After becoming exposed, each agent waits a random number of days before becoming infected. Similarly, after becoming infected, each agent waits a random number of days before becoming recovered. We used lognormal distributions, with parameters consistent with current estimates [21-26], to determine these durations. We also verified that our conclusions were insensitive to small changes in disease progression parameter estimates by running simulations using the defaults specified in another agent-based model[26]. Recovered agents do not become susceptible within the timeframe of our simulations (100 days). To initialize the model, we assumed each student has a 0.1% chance of being initially exposed and a 0.1% chance of being initially infected. In a student population of 5000, these probabilities yield an average of 5 infected individuals and 5 exposed individuals upon return to campus. We also ran all reported simulations with each student having a 0.5% chance of being initially exposed and a 0.5% chance of being initially infected, and we observed very similar results under this less optimistic initial condition (See eFigure 1, eFigure 2, eTable 1, and eTable 2).

In addition to the SEIR dynamics, we also explicitly modelled the impact of various mitigation strategies on the agents. In simulations with surveillance testing, we modelled weekly surveillance tests with 7 days between tests for each agent and the entire population tested every 7 days. For pooled surveillance we pooled agents into groups of size 5 and positive pools were followed by individual tests for each agent in the pool. Pooled and individual tests were assumed to have a uniform delay of 1 day between the test administration and the test result, with a 1.0% false negative rate and a 0.1% false positive rate. Upon receipt of a positive test result, an agent is moved to isolation for 10 days and no longer contributes to any interactions. In simulations with contact tracing, each positive test leads to a random number (possibly 0) of agents being flagged as contacts. Thus, each day, a random number of agents were selected to be reported as contacts of the agents whose positive test results were returned that day. Approximately 15% of the number of reported contacts (or the total number of unisolated, exposed and infected agents, whichever is smaller) were selected from the current pool of unisolated infected or exposed agents, while the remaining number of reported contacts were selected from among the susceptible or recovered agents. The number of contacts reported per positive test was determined by an empirical distribution based on 2470 contacts reported from 971 positive samples collected by Duke University’s contact tracing program during the 2020-2021 academic year (See Supplemental Data). The 15% contact tracing effectiveness was estimated using the results of surveillance testing of quarantined contacts at Duke University. This simple framework allows us to incorporate both the actual number of contacts reported per positive case, and the fraction of reported contacts that were not actually exposed.

For a visual overview of the model architecture see eFigure 3, and for specific model parameter choices used in this study see eTable 3.

## Study Design

In order to assess the impact of various mitigation strategies in the face of an uncertain and potentially evolving environment in Fall 2021, we performed numerical simulations of our model under several environmental conditions and in combination with several mitigation strategies. To give a complete description of an environmental condition, we began with our baseline model parameter values and then we considered deviations of the following parameters: the vaccine effectiveness (90%, 75%, and 50%), the interaction multiplier (1, 10, 20), and the outside community prevalence (0.1% and 1.0%). We considered deviations in the vaccine effectiveness to account for the uncertainty introduced by the possibility of variants and waning immunity. We varied the interaction multiplier in an effort to account for the uncertainty arising from increased density on campus relative to the 2020-2021 academic year and the likelihood of increased numbers of unmasked, indoor interactions both inside and outside the classroom relative to the 2020-2021 academic year [1]. A multiplier of M indicates M times as many interactions as estimated during the Spring semester of 2021 when mitigations such as masking and social distancing were strictly enforced. We considered two values of outside community prevalence to address the uncertainty around this quantity, because it remains difficult to predict future region-specific prevalence given uncertainty around vaccine uptake levels and potential seasonal effects into the Fall.

For each environmental condition, we ran simulations with one of each of the following four mitigation strategies: no mitigation, surveillance testing and isolation, surveillance testing and isolation in combination with contact tracing and quarantining, and surveillance testing in combination with contact tracing and targeted testing of reported contacts every 2 days for 10 days. In the latter model, exposed contacts were not isolated. For each choice of environmental condition and mitigation strategy, we performed 100 simulations of randomly initialized populations, with the population seeded with an average of either 0.2% or 1.0% of agents exposed and infected on day 1. We report the medians and the 2.75%-97.5% quantile ranges of daily percentages in figures and the medians and interquartile ranges in tables across the 100 simulations of each model.

## Results

Regardless of prevalence levels in the off-campus community, at 90% vaccine effectiveness weekly surveillance testing provides only a modest reduction in campus infections, in most conditions reducing the median cumulative infections by less than 1 percentage point (Figure 1 and Table 1). A reduction in median cumulative prevalence by approximately 5 percentage points is observed at 90% vaccine effectiveness only in the most extreme case of 20-fold increase in human interactions and 1% outside community prevalence (Figure 1 and Table 1).

**Figure 1.**
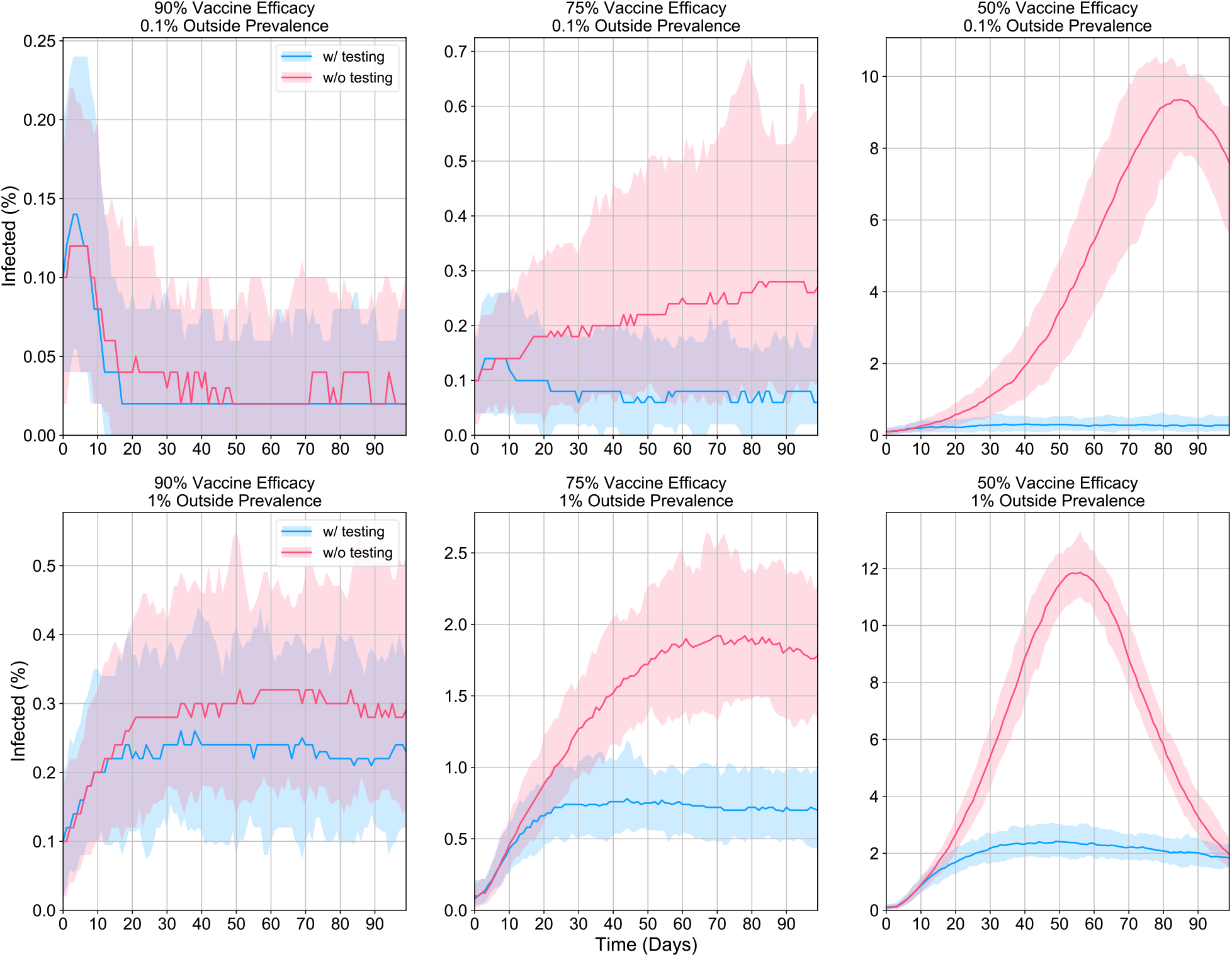
Daily infection prevalence, with and without surveillance testing. The medians (dark lines) and 2.75%-97.5% quantile ranges (light bands) of the daily infection prevalence in the modelled population of 5000 individuals over 100 simulations assuming an interaction multiplier of 10. The outside community infection prevalence was fixed at 0.1% (top) or 1.0% (bottom), with vaccine effectiveness chosen to be 90% (left), 75% (middle) or 50% (right). Simulations were initialized with 0.1% (5) expected initial exposures and 0.1% (5) expected initial infections. All other model parameters were identical across simulations.

**Table 1.** Cumulative infection prevalence across conditions and mitigations. The medians and interquartile ranges of the fraction of cumulative infections in a modelled population of 5000 individuals over 100 simulations of each choice of population parameters and mitigation strategy. For mitigation strategies involving contact tracing, the contact tracing efficacy was fixed at 15%. Simulations were initialized with 0.1% (5) expected initial exposures and 0.1% (5) expected initial infections.

| Population Parameters |  |  | Mitigation Strategies |  |  |  |
| --- | --- | --- | --- | --- | --- | --- |
| Vaccine Effectiveness | Interaction Multiplier | Outside Community Prevalence | None | Surveillance Only | Surveillance & Quarantining Contacts | Surveillance & Testing Contacts |
| 90% | 1 | 0.1% | 0.2% ± 0.1% | 0.2% ± 0.1% | 0.2% ± 0.1% | 0.2% ± 0.1% |
|  |  | 1.0% | 0.4% ± 0.1% | 0.4% ± 0.1% | 0.5% ± 0.1% | 0.4% ± 0.1% |
|  | 10 | 0.1% | 0.6% ± 0.2% | 0.5% ± 0.2% | 0.5% ± 0.1% | 0.5% ± 0.2% |
|  |  | 1.0% | 3.6% ± 0.5% | 2.9% ± 0.3% | 2.9% ± 0.3% | 2.9% ± 0.3% |
|  | 20 | 0.1% | 1.6% ± 0.5% | 0.9% ± 0.2% | 0.9% ± 0.2% | 0.8% ± 0.2% |
|  |  | 1.0% | 11.2% ± 1.3% | 6.4% ± 0.6% | 6.1% ± 0.6% | 6.2% ± 0.5% |
| 75% | 1 | 0.1% | 0.3% ± 0.1% | 0.3% ± 0.1% | 0.3% ± 0.1% | 0.3% ± 0.1% |
|  |  | 1.0% | 0.8% ± 0.2% | 0.8% ± 0.2% | 0.8% ± 0.2% | 0.8% ± 0.2% |
|  | 10 | 0.1% | 2.8% ± 1.1% | 1.2% ± 0.3% | 1.1% ± 0.3% | 1.1% ± 0.2% |
|  |  | 1.0% | 18.5% ± 2.0% | 8.6% ± 0.6% | 8.1% ± 0.7% | 8.0% ± 0.7% |
|  | 20 | 0.1% | 55.8% ± 6.1% | 3.6% ± 1.0% | 3.0% ± 0.8% | 2.7% ± 0.6% |
|  |  | 1.0% | 74.9% ± 1.4% | 24.5% ± 1.8% | 21.1% ± 1.5% | 20.1% ± 1.3% |
| 50% | 1 | 0.1% | 0.4% ± 0.1% | 0.4% ± 0.1% | 0.3% ± 0.1% | 0.3% ± 0.1% |
|  |  | 1.0% | 1.6% ± 0.2% | 1.5% ± 0.2% | 1.5% ± 0.2% | 1.5% ± 0.3% |
|  | 10 | 0.1% | 55.8% ± 6.0% | 3.6% ± 1.0% | 3.0% ± 0.8% | 2.7% ± 0.6% |
|  |  | 1.0% | 75.0% ± 1.4% | 24.5% ± 1.6% | 21.2% ± 1.5% | 20.2% ± 1.1% |
|  | 20 | 0.1% | 96.6% ± 0.4% | 44.8% ± 7.3% | 26.5% ± 5.7% | 20.1% ± 5.1% |
|  |  | 1.0% | 97.4% ± 0.3% | 74.3% ± 2.0% | 61.7% ± 2.2% | 57.6% ± 2.1% |

Infection dynamics are different in conditions where vaccine effectiveness dips to 75% or 50%. At 75% vaccine effectiveness, weekly surveillance testing quickly brings the initial pulse of infections to the baseline, while in the no testing scenario, infection numbers rise slowly over the semester and appear to not reach baseline during the semester (Figure 1). In this scenario, surveillance testing only reduces the number of infections by (2-3)-fold over a no testing regime (Figure 1, Table 1, eTable1). If vaccine effectiveness drops to 50%, then weekly surveillance testing is even more important for limiting the spread of infections. With a 10-fold increase in interactions, surveillance testing massively reduces cumulative infections and peak daily infections, while in the absence of testing daily infections peak between 8% and 12% of the population and then slowly drop towards baseline levels as most of the campus population has experienced an infection and is assumed to be protected from reinfection for the duration of the semester (Figure 1 and Table 1).

We next asked whether contact tracing and quarantine helped limit viral transmission in these conditions and compared the effect of quarantine to a regime in which students who were reported as a contact of a positive case would be tested with increased frequency, rather than quarantined. The median and ranges of model predictions are shown in Figure 2 and eFigure2. Model simulations indicated that quarantine does not substantially reduce infection numbers at 90% vaccine effectiveness and only marginally reduces infections over the course of the semester at 75% vaccine effectiveness (Figure 2, eFigure2, Table 1 and eTable1). Quarantine appears to only have a substantial impact on infection numbers at a vaccine effectiveness of 50% and a 20-fold increase in contacts. Surprisingly, in this scenario testing every two days is a more effective strategy than quarantining reported contacts. Across the conditions, increased testing of contacts appears to be at least as effective as quarantine in limiting the spread of infections (Table 1 and eTable1).

**Figure 2.**
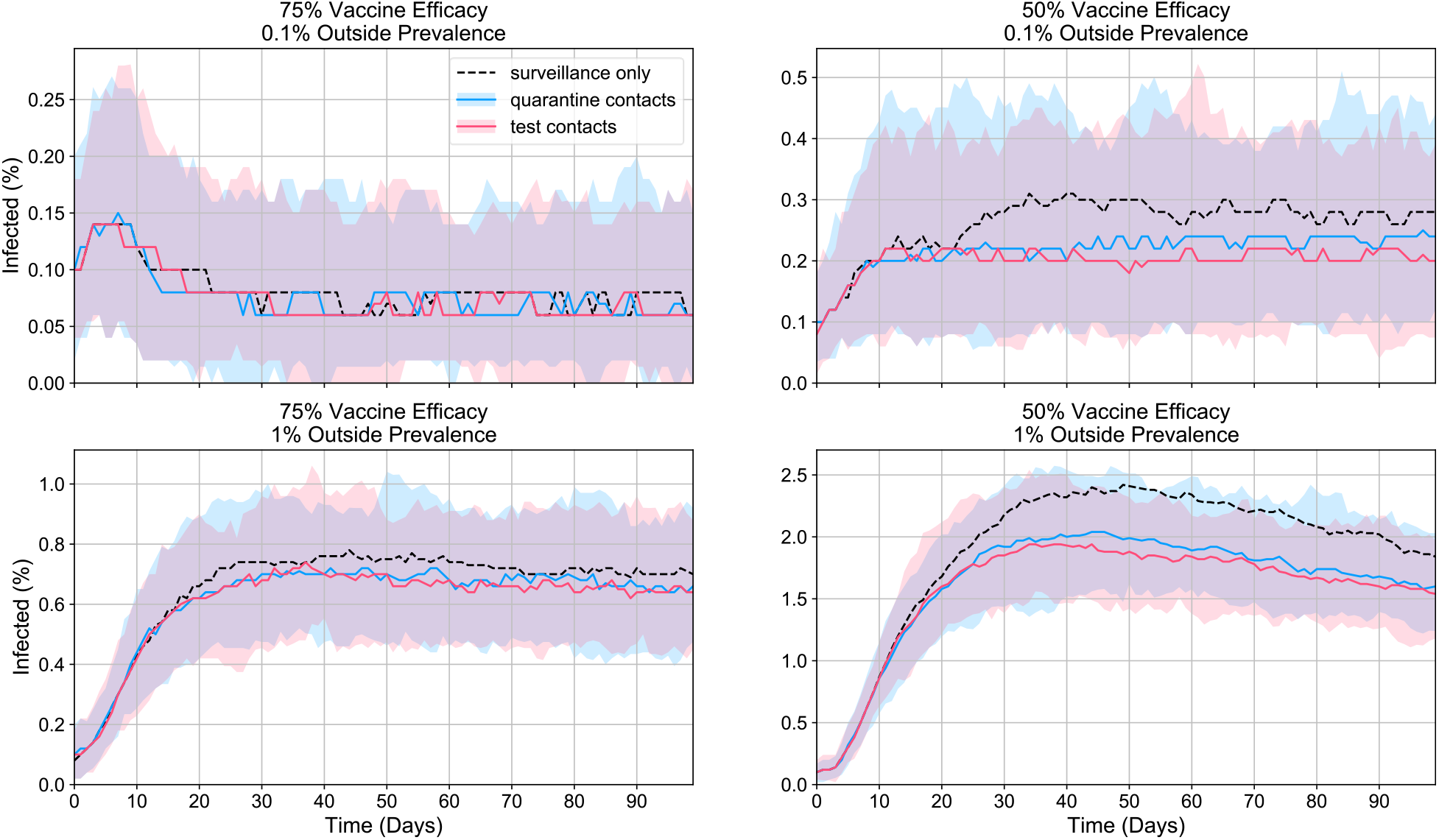
Daily infection prevalence, quarantining vs. testing contacts. The medians (dark lines) and 2.75%-97.5% quantile ranges (light bands) of the daily infection prevalence in a modelled population of 5000 individuals over 100 simulations assuming an interaction multiplier of 10 and either surveillance testing only, surveillance testing with quarantining of contacts, or surveillance testing with testing of contacts every 2 days. The outside community infection prevalence was fixed at 0.1% (top) or 1.0% (bottom), with vaccine effectiveness chosen to be 75% (left) or 50% (right). Contact tracing efficacy was fixed at 15%. Simulations were initialized with 0.1% (5) expected initial exposures and 0.1% (5) expected initial infections. All other model parameters were identical across simulations.

For planning purposes, it is important for universities to estimate the number of rooms needed for quarantine and isolation. Using estimates from the analyses above, we calculated the maximum number of daily isolations and quarantines as a percentage of the total population assuming a 10-day isolation or quarantine period. With surveillance testing and quarantine, the median maximum number in isolation or quarantine ranged between 0.4% (20 for a population of 5,000) to 28.2% (1,410) (Table 2). For the 90% effectiveness scenario, the maximum isolated and quarantine are likely needed at the very beginning of the semester where the peak of infections occur, and for the 75% and 50% scenarios, the peak of infections occurs more towards mid- or late-semester (Figure 1).

As an alternative to quarantine, we examined the outcome of testing contact-traced students every 2 days. Model outputs indicate that testing these students every 2 days has about the same impact on the number of infections over the course of the semester across all scenarios, suggesting that this increased testing cadence may be a viable alternative to quarantine on campuses with 100% of students vaccinated. A benefit of increased testing over quarantine is a substantial reduction in the maximum and total number of students in isolation and quarantine (Table 2 and eTable 2), thus conserving considerable space and human resources needed for quarantine, and more critically allowing students to continue to fully engage in the campus learning environment.

**Table 2.** Maximum fraction of isolated and quarantined individuals. The medians and interquartile ranges of the daily maximum fraction of agents in isolation or quarantine in a modelled population of 5000 individuals for 100 simulations of each choice of population parameters and mitigation strategies. For mitigation strategies involving contact tracing, the contact tracing efficacy was fixed at 15%. Simulations were initialized with 0.1% (5) expected initial exposures and 0.1% (5) expected initial infections.

| Population Parameters |  |  | Mitigation Strategies |  |  |
| --- | --- | --- | --- | --- | --- |
| Vaccine Effectiveness | Interaction Multiplier | Outside Community Prevalence | Surveillance Only | Surveillance & Quarantining Contacts | Surveillance & Testing Contacts |
| 90% | 1 | 0.1% | 0.1% ± 0.1% | 0.4% ± 0.2% | 0.1% ± 0.1% |
| | | 1.0% | 0.1% ± 0.1% | 0.5% ± 0.2% | 0.1% ± 0.1% |
| | 10 | 0.1% | 0.2% ± 0.1% | 0.5% ± 0.2% | 0.2% ± 0.1% |
| | | 1.0% | 0.4% ± 0.1% | 1.3% ± 0.2% | 0.5% ± 0.1% |
| | 20 | 0.1% | 0.2% ± 0.1% | 0.6% ± 0.2% | 0.2% ± 0.1% |
| | | 1.0% | 0.9% ± 0.1% | 2.5% ± 0.4% | 0.9% ± 0.1% |
| 75% | 1 | 0.1% | 0.1% ± 0.1% | 0.4% ± 0.2% | 0.1% ± 0.1% |
| | | 1.0% | 0.2% ± 0.1% | 0.5% ± 0.2% | 0.2% ± 0.1% |
| | 10 | 0.1% | 0.2% ± 0.1% | 0.7% ± 0.3% | 0.2% ± 0.1% |
| | | 1.0% | 1.1% ± 0.2% | 3.3% ± 0.6% | 1.1% ± 0.2% |
| | 20 | 0.1% | 0.6% ± 0.2% | 1.5% ± 0.5% | 0.5% ± 0.1% |
| | | 1.0% | 3.2% ± 0.4% | 7.7% ± 0.9% | 2.7% ± 0.3% |
| 50% | 1 | 0.1% | 0.2% ± 0.1% | 0.4% ± 0.2% | 0.1% ± 0.1% |
| | | 1.0% | 0.3% ± 0.1% | 0.8% ± 0.2% | 0.3% ± 0.0% |
| | 10 | 0.1% | 0.6% ± 0.2% | 1.5% ± 0.5% | 0.5% ± 0.1% |
| | | 1.0% | 3.2% ± 0.4% | 7.8% ± 1.0% | 2.7% ± 0.3% |
| | 20 | 0.1% | 7.8% ± 1.4% | 12.0% ± 3.2% | 3.3% ± 0.8% |
| | | 1.0% | 14.7% ± 1.2% | 28.2% ± 2.8% | 10.3% ± 1.0% |

## Discussion

There is a growing need to re-evaluate the importance of university COVID-19 protocols the availability of effective vaccines. Most universities are encouraging vaccination, and many have mandated SARS-CoV-2 vaccination for all students. Recent guidance from the CDC suggests that mitigations like testing, quarantine, and masking are not necessary for vaccinated populations [1], and universities are likely to adopt these guidelines as university policy. The relaxation of mitigations will necessarily increase the number of interactions within student populations, and thus, without surveillance testing, limiting infections on campus will rely almost entirely on the capability of vaccines to prevent infection and transmission.

Starting with the assumption that 100% of the student population will be vaccinated, we found that in the most optimistic circumstances where vaccines reduce the probability of transmission by 90%, infections are low and even a surge of infections quickly decays in the absence of surveillance testing (Figure 1 and Table 1). However, if vaccine effectiveness drops to 75%, infection dynamics changed and a substantial number of on-campus infections can be observed in the absence of testing. At 50% vaccine effectiveness, the model predicts the possibility of on-campus community spread and exponential growth of infections that could cause substantial outbreaks. At both 75% and 50% effectiveness, surveillance testing alone causes a marked reduction in infections (Figure 1 and eFigure1, Table 1 and eTable1). Our results are qualitatively consistent with previous modeling work pertaining to university populations in the Fall 2021 semester [27]. Indeed, under somewhat different models and parameters than those considered here, they also find that “less effective vaccines or incidence of new variants may require additional intervention such as screening testing” for universities to reopen safely.

Vaccine effectiveness could be reduced to lower levels by a number of mechanisms. First, while BNT162b2 and mRNA-1273 vaccines are reported to be 90% efficient at blocking infection and transmission [3-5], the Ad26.COV2.S vaccine efficacy is considerably lower at 67% [6]. Thus, a high percentage of individuals with the Ad26.COV2.S vaccine on campus could push the average effectiveness below 75%. Second, it is not yet known how long vaccine-induced immunity will last. It is likely that it will diminish slowly over some time frame, and that would lead to a drop in vaccine effectiveness over time. Finally, several genetic variants exhibit some resistance to vaccine-induced neutralizing antibodies [7-10], and while our current vaccines are reported to be effective against variants, they may not have maximum effectiveness [28, 29]. As populations have become partially vaccinated, there is selective pressure favoring variants that can bypass immunity, so the possibility of newly emerging variants with the capacity to bypass some immunity may be an issue in the future.

In addition to vaccine effectiveness, we identified other factors that impact on-campus infection rates. If interactions increase as expected as mitigations such as masking are relaxed both outside the classroom and inside, we observed that the increased interactions result in higher infection rates at all vaccine efficiencies (Table 1 and eTable 1). However, it is hard to predict with any precision how much interactions will increase, so it is not clear how relevant the choices of interaction levels were. We also investigated how off-campus community prevalence impacted on-campus infections and found that increased off-campus prevalence drives an increase in campus infection rates. Off-campus prevalence rates are difficult to predict for the fall semester and will likely depend on the percentage of the off-campus population that is vaccinated. Thus, universities may be differentially impacted based on local vaccination uptake rates.

In addition to surveillance testing, we investigated the role of quarantine across the same scenarios (Figure 2 and eFigure 2). Our findings indicate that quarantine has little or no impact on campus infection rates in most scenarios and only has substantial impacts in scenarios where vaccine effectiveness drops to 50% (Figure 2 and Table 1). We also asked whether an increased testing cadence (every other day) for reported contacts would be as efficient as quarantine in preventing new infections. Model simulations indicate that across scenarios, testing every other day was as efficient as quarantine. These findings support the suggestion that quarantine may be unnecessary for a well-vaccinated student population.

Modeling approaches such as the one used here are subject to a variety of limitations, many related to parameter estimation and assumptions about model structure. Although we expect that the findings will be generally applicable to universities, important parameters in the model were fit to infection dynamics data collected by the surveillance program at Duke University during the 2020-2021 academic year, and therefore outcomes may be context specific. In particular, we used a parameter fitting procedure to estimate the number of daily interactions for our model. We also used the empirical contact tracing distribution and contact tracing efficiency observed by Duke University during the 2020-2021 academic year. We note that the accuracy of self-reporting is unknown. Moreover, in the absence of information on interaction networks, the model assumes homogeneous mixing. Finally, the model only tracks infection dynamics and does not account for vaccine effects on the course of the disease. If disease severity is diminished to acceptable levels in vaccinated individuals [4, 5], the tolerance for infections on campus may be increased, as the outcome of these infections will be less impactful.

A further limitation of our study stems from uncertainty about the impact vaccines will have on the course of infection in the modelled population. Rather than modify disease progression parameters (which specify the distributions for the time from exposure to infection and from infection to recovery) for a vaccinated population, our model directly modifies transmission probabilities so that a vaccine effectiveness of X% is expected to prevent that percentage of potential exposures. This is not the only possible model choice that could be used to explain observed vaccine efficacies, but it is consistent with emerging evidence that vaccinated individuals can become infected [11, 30, 31]. Furthermore, this study only models a fully vaccinated student population and so only directly models universities with a vaccine mandate. That said, we expect the total number of infections over the semester to increase monotonically with the fraction of the population that is unvaccinated. It is not yet clear whether infected individuals who are vaccinated will systematically have lower viral loads and thus be less likely to transmit [32, 33]. If true, then infection rates may be lower than what the model predicts.

Aside from limiting viral transmission on campus, surveillance testing provides other useful public health information. Sequencing of surveillance positives has been used to track the arrival or expansion of particular variants that have attributes such as increased transmissibility or disease severity and enhanced resistance to neutralizing antibodies. The constitution of variants in the population could inform policies on mitigations. Moreover, as policies on gatherings, masking and other mitigations are relaxed, surveillance testing gives real-time feedback on whether relaxations are triggering unwanted spread of SARS-CoV-2 so that timely adjustments can be made.

## Conclusion

Our results only support the elimination of surveillance testing in the most optimistic conditions. In the face of potentially decreasing vaccine effectiveness due to variants or waning immunity, our findings indicate an important role for continued surveillance even on campuses with fully vaccinated student populations. Our findings also indicate that quarantine has only a minimal impact in preventing new infections in most scenarios, and where it does have an impact, it can be replaced by an increased testing cadence for exposed individuals.

## Data Availability

The data, model, and scripts used to perform the analyses and generate the figures and tables presented in this study are available at https://gitlab.com/duke-covid-modeling/covid_campus_interventions_public

## Supplemental Figures and Tables

**eFigure 1.**
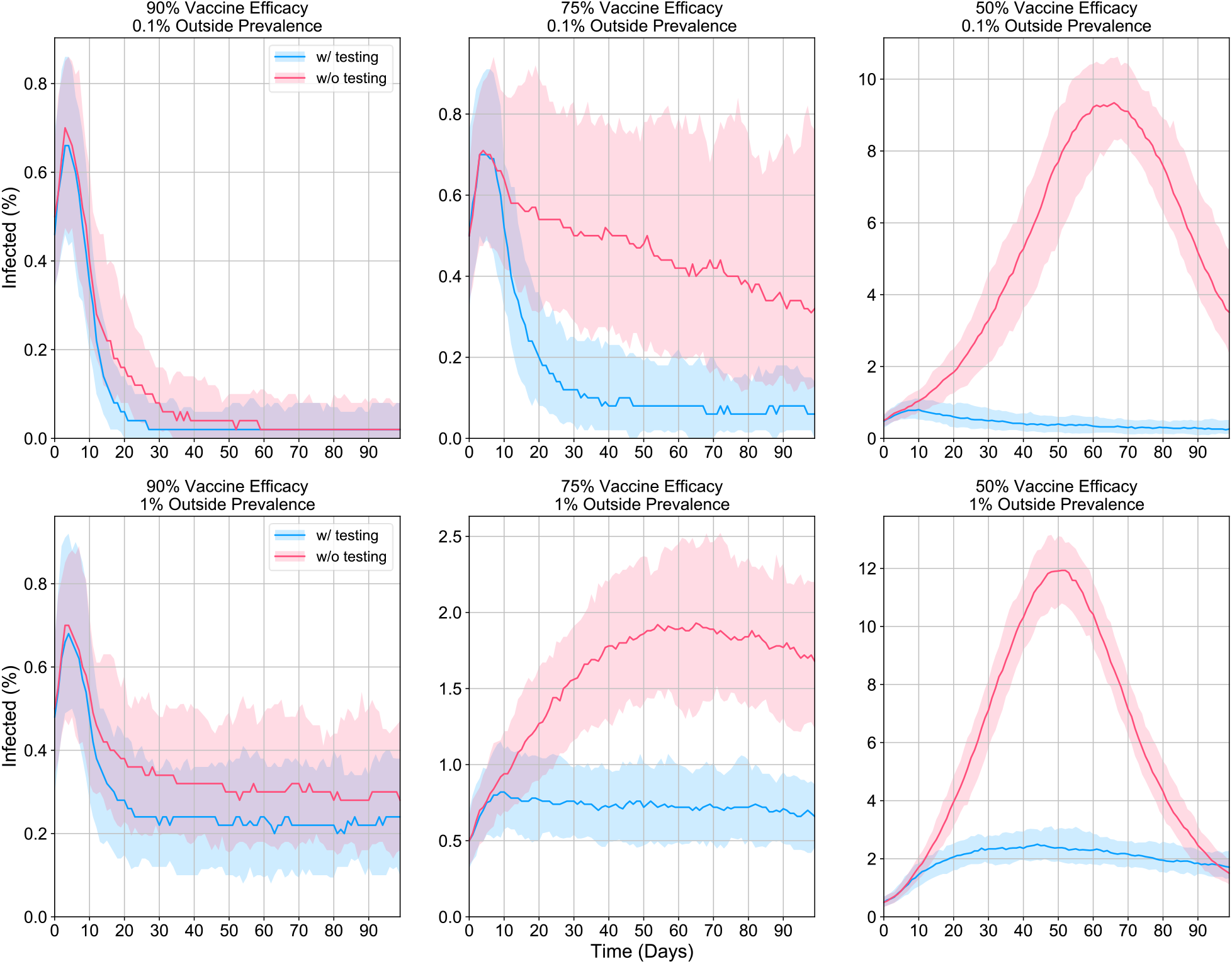
Daily infection prevalence, with and without surveillance testing. The medians (dark lines) and 2.75%-97.5% quantile ranges (light bands) of the daily infection prevalence in the modelled population of 5000 individuals over 100 simulations assuming an interaction multiplier of 10. The outside community infection prevalence was fixed at 0.1% (top) or 1.0% (bottom), with vaccine effectiveness chosen to be 90% (left), 75% (middle) or 50% (right). Simulations were initialized with 0.5% (25) expected initial exposures and 0.5% (25) expected initial infections. All other model parameters were identical across simulations.

**eFigure 2.**
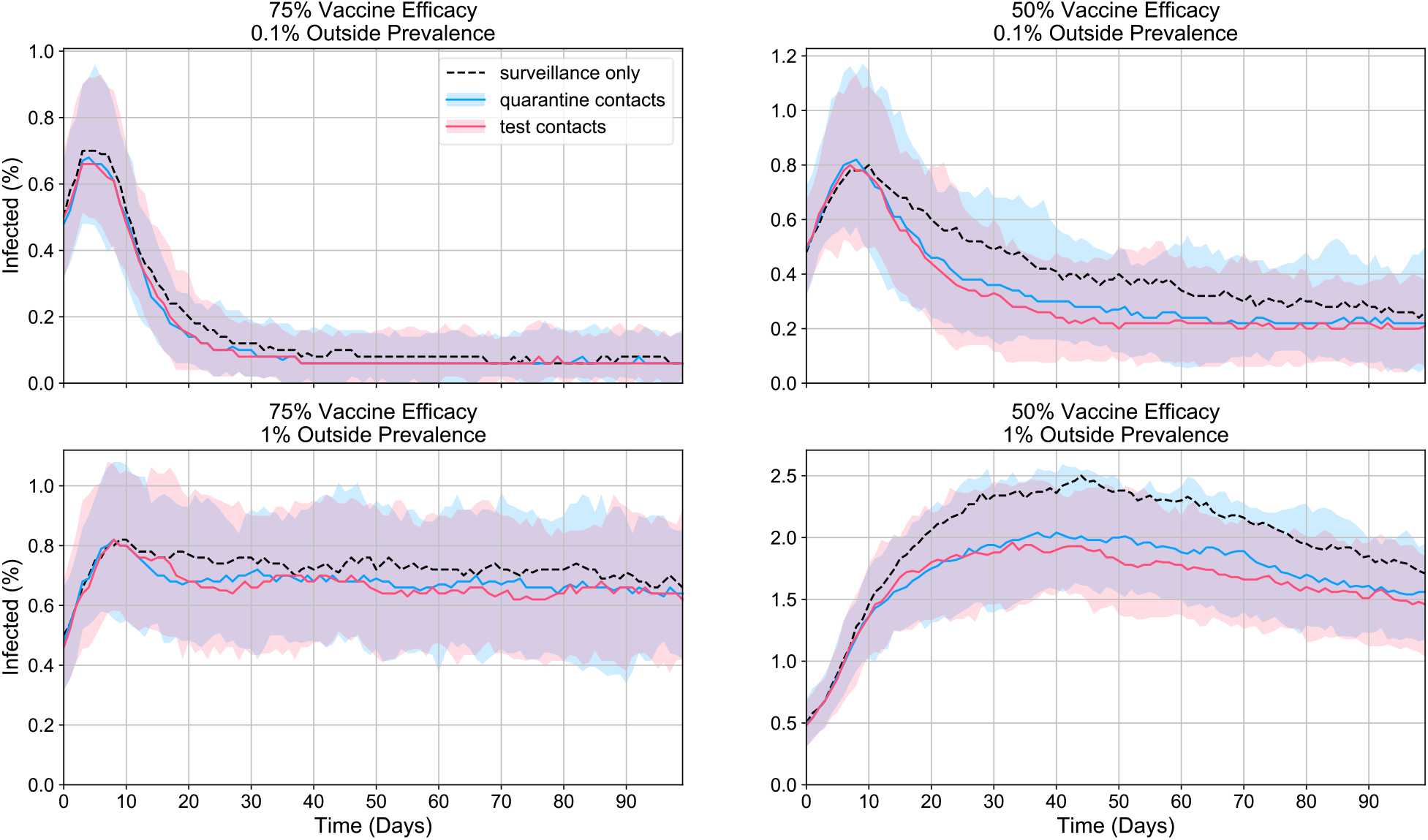
Daily infection prevalence, quarantining vs. testing contacts. The medians (dark lines) and 2.75%-97.5% quantile ranges (light bands) of the daily infection prevalence in a modelled population of 5000 individuals over 100 simulations assuming an interaction multiplier of 10 and either surveillance testing only, surveillance testing with quarantining of contacts, or surveillance testing with testing of contacts every 2 days. The outside community infection prevalence was fixed at 0.1% (top) or 1.0% (bottom), with vaccine effectiveness chosen to be 75% (left) or 50% (right). Contact tracing efficacy was fixed at 15%. Simulations were initialized with 0.5% (25) expected initial exposures and 0.5% (25) expected initial infections. All other model parameters were identical across simulations.

**eFigure 3.**
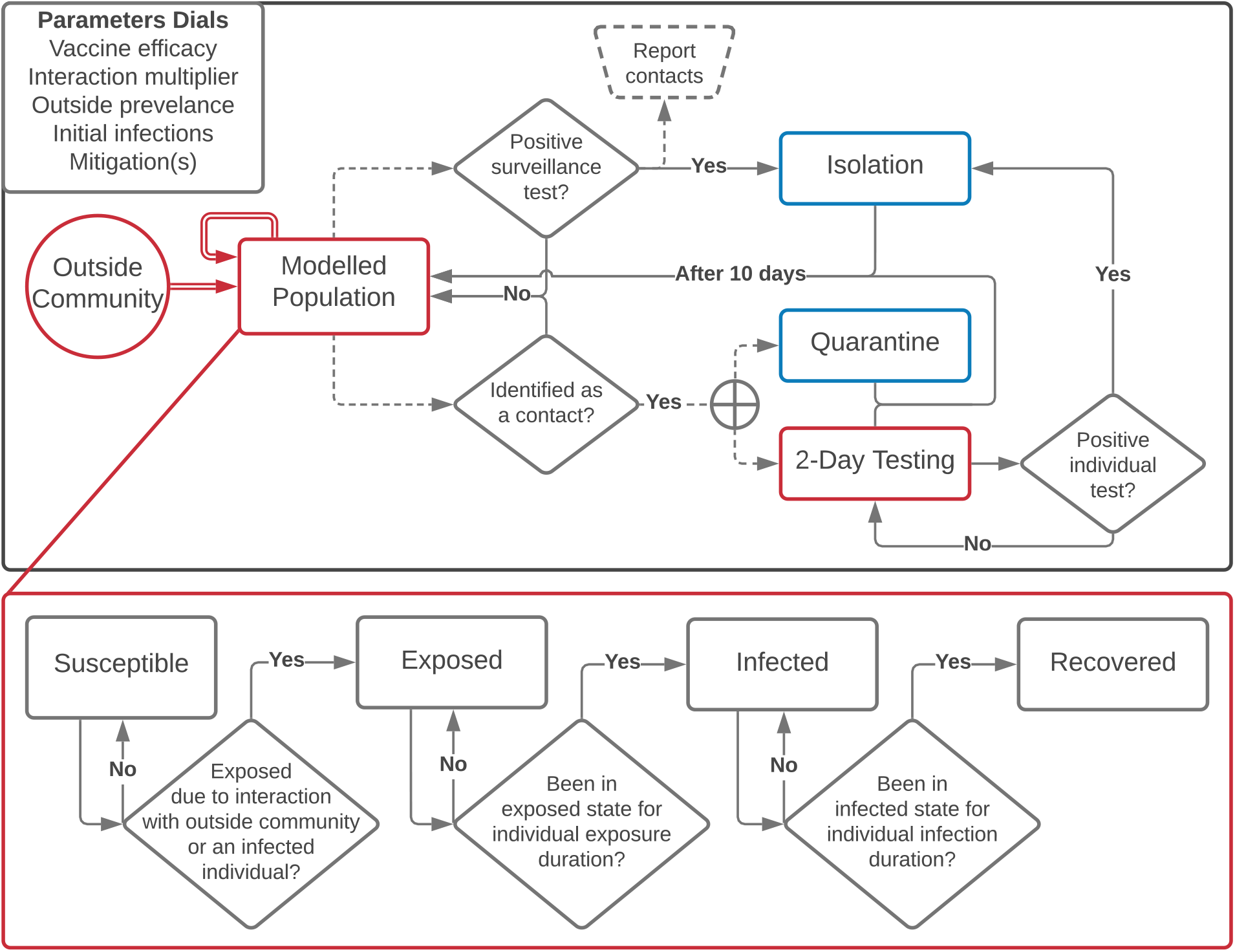
Overview of model architecture. (Top) Flowchart describing each of the four mitigation strategies considered in this study: 1) no testing, 2) surveillance testing only, 3) surveillance with quarantining of contacts, and 4) surveillance with follow-up testing of contacts every 2 days. Dashed arrows indicate optional pathways if the specified mitigation is being modelled. Individuals in red boxes may have interactions with others in the modelled population, while individuals with blue attributes are isolated from the modelled population and outside community. (Bottom) Flowchart of SEIR model and agent health state progression. Arrows indicate transitions between states and daily update rules.

**eTable 1.** Cumulative infection prevalence across conditions and mitigations. The medians and interquartile ranges of the fraction of cumulative infections in a modelled population of 5000 individuals over 100 simulations of each choice of population parameters and mitigation strategy. For mitigation strategies involving contact tracing, the contact tracing efficacy was fixed at 15%. Simulations were initialized with 0.5% (25) expected initial exposures and 0.5% (25) expected initial infections.

| Population Parameters |  |  | Mitigation Strategies |  |  |  |
| --- | --- | --- | --- | --- | --- | --- |
| Vaccine Effectiveness | Interaction Multiplier | Outside Community Prevalence | None | Surveillance Only | Surveillance & Quarantining Contacts | Surveillance & Testing Contacts |
| 90% | 1 | 0.1% | 1.0% ± 0.2% | 1.0% ± 0.2% | 1.0% ± 0.2% | 1.0% ± 0.2% |
|  |  | 1.0% | 1.3% ± 0.2% | 1.2% ± 0.2% | 1.2% ± 0.2% | 1.2% ± 0.2% |
|  | 10 | 0.1% | 1.7% ± 0.3% | 1.4% ± 0.2% | 1.4% ± 0.3% | 1.4% ± 0.2% |
|  |  | 1.0% | 4.8% ± 0.6% | 3.8% ± 0.4% | 3.8% ± 0.4% | 3.8% ± 0.3% |
|  | 20 | 0.1% | 3.7% ± 0.9% | 1.9% ± 0.4% | 1.9% ± 0.3% | 1.9% ± 0.2% |
|  |  | 1.0% | 13.0% ± 1.4% | 7.4% ± 0.7% | 7.0% ± 0.5% | 7.1% ± 0.8% |
| 75% | 1 | 0.1% | 1.1% ± 0.2% | 1.1% ± 0.2% | 1.1% ± 0.2% | 1.1% ± 0.2% |
|  |  | 1.0% | 1.7% ± 0.3% | 1.6% ± 0.3% | 1.6% ± 0.3% | 1.6% ± 0.2% |
|  | 10 | 0.1% | 6.3% ± 1.4% | 2.4% ± 0.5% | 2.1% ± 0.4% | 2.1% ± 0.3% |
|  |  | 1.0% | 20.7% ± 2.0% | 9.6% ± 0.7% | 9.0% ± 0.8% | 8.8% ± 0.8% |
|  | 20 | 0.1% | 66.5% ± 2.6% | 5.8% ± 1.0% | 4.7% ± 1.0% | 4.5% ± 0.8% |
|  |  | 1.0% | 75.7% ± 1.4% | 26.1% ± 1.9% | 22.1% ± 1.5% | 21.2% ± 1.5% |
| 50% | 1 | 0.1% | 1.3% ± 0.2% | 1.2% ± 0.2% | 1.2% ± 0.2% | 1.2% ± 0.2% |
|  |  | 1.0% | 2.6% ± 0.3% | 2.3% ± 0.4% | 2.3% ± 0.3% | 2.3% ± 0.3% |
|  | 10 | 0.1% | 66.5% ± 2.7% | 5.8% ± 1.0% | 4.7% ± 1.0% | 4.5% ± 0.8% |
|  |  | 1.0% | 75.8% ± 1.4% | 25.7% ± 2.0% | 22.0% ± 1.5% | 21.2% ± 1.4% |
|  | 20 | 0.1% | 96.7% ± 0.4% | 54.3% ± 5.6% | 33.7% ± 6.0% | 25.4% ± 5.1% |
|  |  | 1.0% | 97.5% ± 0.4% | 74.3% ± 1.5% | 62.4% ± 2.5% | 58.7% ± 2.3% |

**eTable 2.** Maximum fraction of isolated and quarantined individuals. The medians and interquartile ranges of the daily maximum fraction of agents in isolation or quarantine in a modelled population of 5000 individuals for 100 simulations of each choice of population parameters and mitigation strategies. For mitigation strategies involving contact tracing, the contact tracing efficacy was fixed at 15%. Simulations were initialized with 0.5% (25) expected initial exposures and 0.5% (25) expected initial infections.

| Population Parameters |  |  | Mitigation Strategies |  |  |
| --- | --- | --- | --- | --- | --- |
| Vaccine Effectiveness | Interaction Multiplier | Outside Community Prevalence | Surveillance Only | Surveillance & Quarantining Contacts | Surveillance & Testing Contacts |
| 90% | 1 | 0.1% | 0.7% ± 0.1% | 2.0% ± 0.6% | 0.7% ± 0.1% |
| | | 1.0% | 0.7% ± 0.2% | 2.1% ± 0.7% | 0.7% ± 0.2% |
| | 10 | 0.1% | 0.7% ± 0.2% | 2.1% ± 0.7% | 0.8% ± 0.2% |
| | | 1.0% | 0.8% ± 0.2% | 2.4% ± 0.6% | 0.9% ± 0.2% |
| | 20 | 0.1% | 0.8% ± 0.2% | 2.2% ± 0.7% | 0.9% ± 0.2% |
| | | 1.0% | 1.0% ± 0.1% | 2.9% ± 0.6% | 1.0% ± 0.2% |
| 75% | 1 | 0.1% | 0.7% ± 0.1% | 2.0% ± 0.5% | 0.7% ± 0.2% |
| | | 1.0% | 0.7% ± 0.2% | 2.1% ± 0.7% | 0.8% ± 0.2% |
| | 10 | 0.1% | 0.9% ± 0.2% | 2.4% ± 0.7% | 0.9% ± 0.2% |
| | | 1.0% | 1.2% ± 0.1% | 3.4% ± 0.5% | 1.2% ± 0.1% |
| | 20 | 0.1% | 1.0% ± 0.2% | 3.1% ± 0.9% | 1.1% ± 0.3% |
| | | 1.0% | 3.3% ± 0.4% | 7.8% ± 0.9% | 2.7% ± 0.3% |
| 50% | 1 | 0.1% | 0.7% ± 0.2% | 2.0% ± 0.5% | 0.8% ± 0.2% |
| | | 1.0% | 0.7% ± 0.2% | 2.1% ± 0.6% | 0.8% ± 0.2% |
| | 10 | 0.1% | 1.0% ± 0.2% | 3.1% ± 0.9% | 1.1% ± 0.3% |
| | | 1.0% | 3.1% ± 0.4% | 7.7% ± 1.0% | 2.7% ± 0.3% |
| | 20 | 0.1% | 8.2% ± 1.4% | 12.9% ± 2.5% | 3.4% ± 0.9% |
| | | 1.0% | 14.5% ± 1.0% | 28.2% ± 2.6% | 10.2% ± 1.0% |

**eTable 3.** Model parameters. The set of model parameters used in this study. Disease progression parameters and reported contacts determined by parameterized distributions: (*) lognormal(mean=4.6, stdev=4.8, minimum=3) or lognormal(mean=4.5, stdev=1.5, minimum=1), (**) 1.1% drawn from lognormal(mean=14.0, stdev=2.4, minimum=1) and 98.9% drawn from lognormal(mean=8.0, stdev=2.0, minimum=1) or lognormal(mean=8.0, stdev=2.0, minimum=1), and (***) the empirical distribution from the Duke University 20-21 academic year with an average of 2.54 contacts per positive test. The function lognormal(mean, stdev, minimum) returns a sample of a log normal distribution, with specified mean and standard deviation, and with values rounded to the nearest integer, with values less than the specified minimum replaced by that minimum.

| Variable | Description | Value |
| --- | --- | --- |
| **Disease Progression** |  |  |
| Duration of exposed | The number of days agents stay in the exposed state. | (*) |
| Duration of infected | The number of days agents stay in the infected state. | (**) |
| **Population** |  |  |
| Size | The number of individuals in the modelled population. | 5000 |
| Initial exposed | The average fraction of individuals initially in the exposed state. | 0.1% / 0.5% |
| Initial infected | The average fraction of individuals initially in the infected state. | 0.1% / 0.5% |
| Baseline daily interactions | The baseline number of unique daily interactions between members of the modelled population on average. | 1.67 |
| Baseline interaction exposure probability | The baseline probability that a susceptible-infected interaction will result in an exposure. | 2.60% |
| Baseline outside daily interactions | The baseline number of unique daily interactions individuals will have with the outside community on average. | 0.995 |
| Baseline outside interaction exposure probability | The baseline probability that an interaction between a susceptible member of the modelled population and the outside community will result in an exposure. | 2.60% |
| Outside prevalence | The current fraction that is infected in the outside community. | 0.1% / 1.0% |
| Interaction multiplier | Factor by which the baseline number of interactions is multiplied. | 1 / 10 / 20 |
| Vaccine effectiveness | Percent reduction in the baseline interaction exposure probability. | 90% / 75% / 50% |
| **Surveillance and Individual Testing** |  |  |
| Days between surveillance tests | The number of days between tests of individual agents, and the number of days required to test the entire population exactly once. | 7 |
| Testing pool size | The number of tests pooled together for initial screening. | 5 |
| Test FPR | The false positive rate of pooled and individual tests. | 0.1% |
| Test FNR | The false negative rate of pooled and individual tests. | 1.0% |
| Test results delay | The number of days until test results become available. | 1 |
| Isolation duration | The number of days a positive case spends in isolation. | 10 |
| **Contact Tracing Parameters** |  |  |
| Contacts reported | The number of contacts an agent will report if they test positive | (***) |
| Tracing effectiveness | The maximum fraction of reported contacts who are exposed or infected at the time of the contact trace. | 15% |
| Quarantine duration | The number of days a reported contact of a positive test spends in quarantine (or undergoes additional testing) | 10 |

